# Early Prediction of Discharge Outcomes in Geriatric Rehabilitation: Development and Temporal Validation of a Multiclass Model

**DOI:** 10.64898/2026.11.10.26367591

**Authors:** Erez Kurzweil, Racheli Hefetz, Chen Yanover, Inbal Galor, Nechama Markovitz, Inbal Goldshtein

## Abstract

**Objective:** To develop and validate a model that predicts on day 7 of subacute rehabilitation (timed to inform the first multidisciplinary team meeting) the probabilities of three mutually exclusive outcomes: minimal dependency, nursing-care dependency, or medical deterioration.

**Design:** Retrospective cohort study with temporal validation. Multinomial regression and XGBoost models were trained on 80% of admissions during the years 2016-2024 of members of the largest national health maintenance organization (HMO), tested on the remaining 20% and members of other HMOs, and validated on year 2025 admissions. Models output patient-level outcome probabilities with the strongest feature contributors.

**Setting:** A public geriatric rehabilitation hospital.

**Participants:** All consecutive first admissions (N=10,920; median age 80 years; median length of stay 35 days).

**Interventions:** Not applicable.

**Main Outcome Measures:** Discharge disposition: minimal dependency, nursing-care dependency, or medical deterioration (transfer to acute care without return, or in-hospital death). Predictors were demographic, functional, cognitive, clinical, and laboratory variables documented before day 7 with linkage to pre-admission community health records. Performance was assessed by multiclass area under the receiver operating characteristic curve (AUC), Brier score and positive predictive value (PPV).

**Results:** Overall, 34% of patients were discharged with minimal dependency, 44% with nursing-care dependency, and 22% deteriorated medically. Both models performed comparably. Multinomial regression achieved an AUC of 0.81 (95% CI: 0.79-0.83) compared with 0.72 (0.71-0.73) for FIM alone, and it was maintained on temporal validation (AUC 0.80, 0.78-0.82). At a ≥60% probability threshold, the model flagged 32% of patients for nursing-care dependency and 5% for deterioration, with PPVs of 72% and 69%, respectively.

**Conclusions:** Day-7 multiclass risk estimation from routinely collected data is feasible, interpretable and stable over time. Alerts could prompt earlier social-work involvement and closer monitoring of patients at risk of deterioration; prospective controlled evaluation is warranted.

## 1. INTRODUCTION

Subacute geriatric rehabilitation serves older adults who lost functional independence after hip fractures, stroke, prolonged ventilation or acute illness. Outcomes diverge: many return home with functional gains, many remain nursing-care dependent, and others deteriorate medically — requiring acute-care transfer, or die during the rehabilitation stay.

These trajectories carry different implications for patients, families, clinical teams, and health systems. Patients heading toward nursing-care dependency often require extensive social work involvement to facilitate access to entitlements and supportive care services. The social worker prepares the family and initiates application for the National Insurance long-term care benefits that fund ongoing home-care assistance. ^1^ Because this process is lengthy and often initiated late, discharge is frequently deferred until the needed support is in place. ^2^ These avoidable prolonged admissions limited rehabilitation capacity without clinical benefit.

Patients at risk of medical deterioration, by contrast, require intensified medical monitoring, proactive specialist consultation, and early goals-of-care discussions. Quantifying the risk of medical deterioration could also support staff allocation across wards. Lastly, patients who achieve successful rehabilitation require discharge planning focused on community reintegration.

Rehabilitation decisions are made at multidisciplinary team meetings; at [blinded hospital], the first convenes about 7 days post-admission and recurs roughly every two weeks. The team reviews each patient’s progress, sets goals, intervention intensity, and anticipated discharge timing and aftercare. Early, accurate estimation of anticipated care needs is a major challenge, yet practical tools are lacking. ^3^

Existing prognostic tools rely predominantly on single-index scores such as the Functional Independence Measure (FIM), with modest discriminative ability. ^4,5^ Most studies frame outcomes as binary (home vs. non-home), conflating institutionalization with acute-care transfer, two outcomes requiring different actions. ^6–8^ Destination is influenced by caregiver availability, living situation, socioeconomic status and patient preferences. ^9,10^ The literature more often addresses who returns home than who deteriorates, and rarely combines all three states in a single early prediction framework. ^7,11^ Few studies leverage pre-admission community records or define implementation pathways.

We aimed to develop and validate a multiclass prediction model for these three outcomes using routine data available by the first rehabilitation week.

## 2. METHODS

### 2.1 Study Design and Setting

This retrospective cohort study used electronic health records of a large Israeli health maintenance organization (HMO) and an affiliated public geriatric rehabilitation hospital. All Israeli residents receive a uniform benefits basket through one of four HMOs with longitudinal electronic records; the largest, covers about half the population. [blinded hospital] is a public rehabilitation hospital in central Israel with orthopedic, neurological, general geriatric deconditioning, and chronic ventilation wards. HMO affiliation enables linkage of rehabilitation records with pre-admission community data. Data were extracted using a secure, de-identified data-sharing platform (MDClone, https://www.mdclone.com). The institutional ethics committee approved the study [number 0631-24-RMC]; informed consent was waived given the retrospective, de-identified design. This study is reported in accordance with the TRIPOD+AI statement. ^12^

### 2.2 Study Population

We extracted all consecutive first admissions for inpatient rehabilitation between January 2016 and November 2025. Patients with missing outcome, discharge date or sex were excluded. No exclusions were applied by age, diagnosis, or rehabilitation type, so the model generalizes across the facility’s full patient spectrum. For model training we used January 2016- November 2024 admissions.

### 2.3 Outcome Definition

The primary outcome was eventual discharge disposition with three categories: (1) medical deterioration, (2) nursing-care dependent discharge, and (3) minimal-dependency.

Medical deterioration was defined as transfer to acute care without return, or death during the rehabilitation stay. These events were grouped because they share predictors (acute illness severity, comorbidity burden) and because the number of in-hospital deaths was insufficient to support a reliable separate class.

For medically stable patients, dependency was classified using the Israeli Ministry of Health (MoH) standardized categories, which group patients by required assistance. ^13^

Patients classified at discharge as ‘nursing-care dependent’, or ‘mentally frail’ under the MoH functional assessment were combined into a single outcome class, hereafter termed ‘Nursing-care dependency’. This captures patients needing substantial or continuous activities of daily living (ADL) assistance (from physical dependency with high-acuity needs or from significant cognitive impairment such as dementia) who did not regain independence and required home-care or institutional placement.

Minimal dependency comprised the MoH categories independent or mild physical deterioration (no or only partial ADL assistance, without medical deterioration).

### 2.4 Landmark Design (Feature Extraction Cutoff)

Predictors were restricted to data documented by day 7 post-admission (the first staff meeting). The outcome was not time-restricted: it reflects the eventual disposition regardless of discharge timing, so the forecast window ranged from days to months. Length of stay, shaped by operational (non-clinical) factors was not modelled.

### 2.5 Potential predictors

Predictors came from two sources:

#### Hospital-based data

First-week routine intake included FIM score, Mini-Mental State Examination (MMSE, including the raw score as well as categorization vs. age/education norms), instrumental activities of daily life (iADL), Beiti Bedside functional test, and confusion assessment method for delirium screening, all assessed by trained occupational therapists, physiotherapists and nurses. ^14^

We also included BMI, pressure wounds, vital signs, self-reported pre-morbid motor FIM, upper and lower limb strength, and hand grip dynamometry. Hand grip was documented only in one of the five rehabilitation wards and was thus imputed for others.

Demographic and administrative variables included age, sex, ward, admission year, residential socioeconomic status and population sector (Arab patients combined with the general category due to small counts).

Admission reason was classified as orthopedic, neurologic, post–mechanical ventilation, or deconditioning using admission diagnoses and ward assignment; automated classification was 89% concordant with manual adjudication by the study clinician (n=5,625). Laboratory values through day 7 (complete blood count, metabolic panel, liver enzymes, albumin, TSH, glucose) were included, along with a laboratory frailty index (FI-Lab),^15,16^ the proportion of abnormal results among available tests, to capture subclinical physiological dysregulation distinguishing medical deterioration from nursing-care dependency. Local reference ranges were used where assays differed from the original Howlett thresholds. The number of first-week laboratory tests was also included.

### Pre-admission community health data

For the HMO members (about 80%), pre-admission community data were available: nursing-care allowance eligibility, housebound status, chronic diagnoses, medication exposures [psychoactive medications, opiates, insulin, antibiotics, medications which require close monitoring (TDM, such as Coumadin etc.), and polypharmacy (categorized as 0-4/5-9/10+)], Charlson Comorbidity Index, ^17^ Elixhauser comorbidity index,^18^ the veterans affairs frailty index,^19^ and prior hospitalizations.

For the remaining 20% (other HMOs), these variables were handled with the missing-indicator approach described below.

Lastly, two binary interaction indicators were derived: “assessable-but-burdened” (available Beiti score with Elixhauser index above the development-cohort median) and “low FI-Lab-high-Elixhauser” (FI-Lab below the median and Elixhauser index above the median).

### 2.6 Clinical Decision Support Framework

We defined two use cases for the team meeting. For nursing-care dependency, an alert triggers early social-worker engagement: preparing the family, drafting the supporting letter required for the National Insurance nursing-care allowance benefit application; and arranging home care (including foreign-caregiver licensing) or long-term-care facility placement. These lengthy steps can then proceed in parallel with rehabilitation rather than delaying discharge. For medical deterioration, an alert prompts intensified monitoring (more frequent physician review, proactive specialist or palliative consultation, and early goals-of-care discussion).

The management team set both thresholds at a predicted probability of ≥60%, balancing sensitivity against false-positive burden (unnecessary social-worker involvement or premature family conversations); the threshold can be lowered to flag more patients where capacity allows.

### 2.7 Risk contributors

We designed per-patient local explanation plots to support the multidisciplinary team meeting, analogous to SHAP and Explainable Boosting Machines plots and compliant with OPTICA. ^20,21^ For each patient, the plot displays the top features contributing to the predicted log-odds of each outcome class vs. minimal-dependency, relative to the average patient in the development cohort. Contributions are not causal effects but reflect how the model reached its prediction for that patient. Clinicians cite such explanations as a prerequisite for using prediction tools. ^22,23^

### 2.8 Statistical analysis

A tidymodels preprocessing recipe was applied: handling rare (<5%) factor levels, one-hot encoding, near-zero-variance removal, and centering/scaling of numeric predictors. For continuous predictors with missing values, we used a missing-indicator approach (median imputation plus a binary flag). This suits systematically missing community data and informative missingness (e.g. unmeasured first-week scales may reflect instability).

Development-cohort medians were applied to impute the 2025 validation cohort. The 2016-2024 HMO members sub-cohort was randomly split into training (80%) and an internal held-out test set (20%); members of other HMOs formed the second held-out test-set (transportability across insurers). The 2025 cohort formed a third, temporal test set, to check whether performance holds as clinical practice and case-mix evolve. ^24^ A benchmark model using FIM alone was fitted through the same workflow.

Sample size adequacy was confirmed using the Riley/Pate framework for multinomial models with pmsampsize R package ^25,26^. A multiclass AUC of 0.80, 100 candidate parameters, and observed class prevalences of 22/34/44% required a training set of at least n=4,100 which was met. The data met the 10-events-per-parameter rule.

We compared multinomial regression, with natural splines and a prespecified age-by-sex interaction, and gradient-boosted trees (XGBoost, 200 trees, six hyperparameters tuned over a 30-point Latin hypercube grid). No class-imbalance resampling or reweighting was applied. Models were evaluated on discrimination (multi-class ROC AUC ^27^) and calibration (multiclass Brier score, intercept and slope) on held-out test sets. Per-class performance was assessed overall and within pre-specified subgroups: admission reason (clinical), sector and socioeconomic strata (fairness). Cumulative positive predictive value (PPV) was computed. Predictor importance from the multinomial model was quantified per outcome class using likelihood-ratio chi-square for removing each predictor (all its spline or dummy terms jointly), scaled relative to the strongest predictor; the Supplement additionally ranks class-specific top contributors by joint Wald statistics.

Time to discharge with nursing-care dependency was analysed by Kaplan-Meier, stratified by predicted class (highest assigned probability), with log-rank testing.

Analyses were performed in R 4.5.1.

### 2.9 Open science

No study protocol was prepared. The study was not registered as development of a clinical prediction model is not subject to mandatory registration.

## 3. RESULTS

### 3.1 Study Population

Of 10,920 patients admitted to [blinded hospital] rehabilitation, a discharge outcome was recorded for 10,624 (97%). After exclusions (Figure S1), we divided the population into a main cohort (2016– November 2024, Table 1+ Table S1), and a temporal test set (December 2024-November 2025, Table S2).

**Table 1.** Baseline characteristics by outcome, main cohort (2016-2024). For more characteristics, see Table S2.

| **Characteristic** | **Minimal-dependency**<br>N = 3,241 | **Medical deterioration**<br>N = 2,088 | **Nursing-care dependent**<br>N = 4,207 |
|---|---|---|---|
| Age, Median (IQR) | 76 (70 – 82) | 81 (73 – 87) | 83 (76 – 88) |
| Sex, n (%) |  |  |  |
| Female | 1,864 (58) | 1,069 (51) | 2,579 (61) |
| Male | 1,377 (42) | 1,019 (49) | 1,628 (39) |
| Strength upper limbs, n (%) |  |  |  |
| 2 | 70 (3.0) | 67 (5.9) | 155 (5.8) |
| 3 | 333 (14) | 370 (33) | 846 (32) |
| 4 | 1,228 (52) | 515 (45) | 1,270 (48) |
| 5 | 730 (31) | 180 (16) | 392 (15) |
| Unknown | 880 | 956 | 1,544 |
| Strength lower limbs, n (%) |  |  |  |
| 2 | 263 (11) | 202 (19) | 469 (18) |
| 3 | 625 (27) | 430 (40) | 1,017 (39) |
| 4 | 1,062 (46) | 363 (33) | 930 (36) |
| 5 | 378 (16) | 91 (8.4) | 198 (7.6) |
| Unknown | 913 | 1,002 | 1,593 |
| Hand grip, Median (IQR) | 17 (13 – 22) | 12 (8– 17) | 12 (8 – 16) |
| Unknown | 2,102 | 1,563 | 2,907 |
| Nursing-care eligibility (hospital data), n (%) | 755 (23) | 761 (36) | 1,965 (47) |
| Nursing-care allowance level (community data), n (%) |  |  |  |
| 0 | 3,012 (93) | 1,769 (85) | 3,699 (88) |
| 1 | 31 (1) | 14 (1) | 23 (1) |
| 2 | 49 (2) | 39 (2) | 55 (1) |
| 3 | 78 (2) | 71 (3) | 107 (3) |
| 4 | 38 (1) | 67 (3) | 121 (3) |
| 5 | 16 (0) | 46 (2) | 82 (2) |
| 6 | 17 (1) | 81 (4) | 119 (3) |
| Alcohol use disorder, n (%) | 86 (3) | 59 (3) | 65 (2) |
| Housebound, n (%) | 50 (2) | 158 (8) | 523 (12) |
| Pre-admission Mobility status, n (%) |  |  |  |
| Yes (Mobile) | 3,082 (95) | 1,662 (80) | 3,238 (77) |
| Housebound | 12 (0) | 60 (3) | 107 (3) |
| Bedridden | 5 (0) | 17 (1) | 34 (1) |
| Mobile with assistance | 142 (4) | 349 (17) | 828 (20) |
| Premorbid motor FIM, Median (IQR) | 79 (76 - 89) | 76 (65 - 80) | 73 (62 - 78) |
| Unknown | 795 | 695 | 1,096 |
| Smoking, n (%) |  |  |  |
| never | 1,301 (40) | 814 (39) | 1,861 (44) |
| current | 324 (10) | 216 (10) | 303 (7) |
| past | 516 (16) | 322 (15) | 504 (12) |
| unknown | 1,100 (34) | 736 (35) | 1,539 (37) |
| Charlson comorbidity index, Median (IQR) | 3 (1 – 5) | 5 (3 – 7) | 4 (2 – 7) |
| VA-Frailty index deficits, Median (IQR) | 10 (7 – 13) | 12 (8 – 16) | 12 (9 – 16) |
| Elixhauser score, Median (IQR) | 10 (2 – 19) | 17 (9 – 27) | 14 (6 – 23) |
| Delirium screening, n (%) |  |  |  |
| No delirium | 1,842 (57) | 1,000 (49) | 2,059 (49) |
|  | N = 3,241 | N = 2,088 | N = 4,207 |
| Delirium | 6 (0) | 57 (3) | 106 (3) |
| Unscreened | 1,393 (43) | 1,031 (49) | 2,042 (49) |
| Delirium (diagnoses), n (%) | 104 (3) | 209 (10) | 424 (10) |
| Population sector, n (%) |  |  |  |
| General Jewish population | 2,646 (82) | 1,788 (86) | 3,512 (83) |
| Ultra-orthodox Jewish | 223 (7) | 108 (5) | 300 (7) |
| Immigrants from the former Soviet Union | 300 (9) | 154 (7) | 289 (7) |
| Arab | 72 (2) | 38 (2) | 106 (3) |
| Year, n (%) |  |  |  |
| 2016 | 404 (12) | 194 (9) | 500 (12) |
| 2017 | 384 (12) | 214 (10) | 575 (14) |
| 2018 | 408 (13) | 210 (10) | 557 (13) |
| 2019 | 401 (12) | 228 (11) | 601 (14) |
| 2020 | 315 (10) | 197 (9) | 437 (10) |
| 2021 | 353 (11) | 232 (11) | 464 (11) |
| 2022 | 340 (10) | 248 (12) | 385 (9) |
| 2023 | 337 (10) | 277 (13) | 352 (8) |
| 2024 | 299 (9) | 288 (14) | 336 (8) |
| BMI, n (%) |  |  |  |
| Normal weight | 1,025 (32) | 791 (38) | 1,684 (40) |
| Obesity I | 626 (19) | 329 (16) | 642 (15) |
| Obesity II-III | 313 (10) | 173 (8) | 261 (6) |
| Overweight | 1,166 (36) | 699 (33) | 1,382 (33) |
| Underweight | 93 (3) | 85 (4) | 222 (5) |
| Unknown | 18 (1) | 11 (1) | 16 (0) |
| Lonely, n (%) | 268 (8.3) | 206 (9.9) | 522 (12) |

The main cohort was median age 80 (IQR 73-86), 58% female, and median length of stay 35 days (IQR 22-49). The primary admission reasons were orthopedic (47%), neurologic (20%), de-conditioning (28%), and ventilation weaning (6%). The outcome distribution was: minimal-dependency 3,241 (34%), nursing-care dependent discharge 4,207 (44%), and medical deterioration 2,088 (22%), of whom n=161 died during the rehabilitation stay and n=1,927 were transferred to acute care without return.

Among those who required social worker letters, the average time from index to first letter was 21 days, increasing from 18 days between 2016-2020 to 26 days between 2021-2024. Of the main cohort (n=9,536), 7,535 (79%) were HMO members with linked community data and 2,001 (21%) were from other HMOs. HMO members were split into training (n=6,031) and internal (n=1,504) sets; members of other HMOs formed an external test-set.

### 3.2 Model Performance

Figure 1 depicts similar performance of the multinomial regression model and XGBoost. On the HMO-members test-set, the multinomial model achieved macro-averaged ROC AUC of 0.81 (95% CI 0.79-0.83), and Brier score of 0.22. The multinomial model was well calibrated for all outcomes while XGBoost overestimated low risks and underestimated high ones (Figure S2).

**Figure 1.**
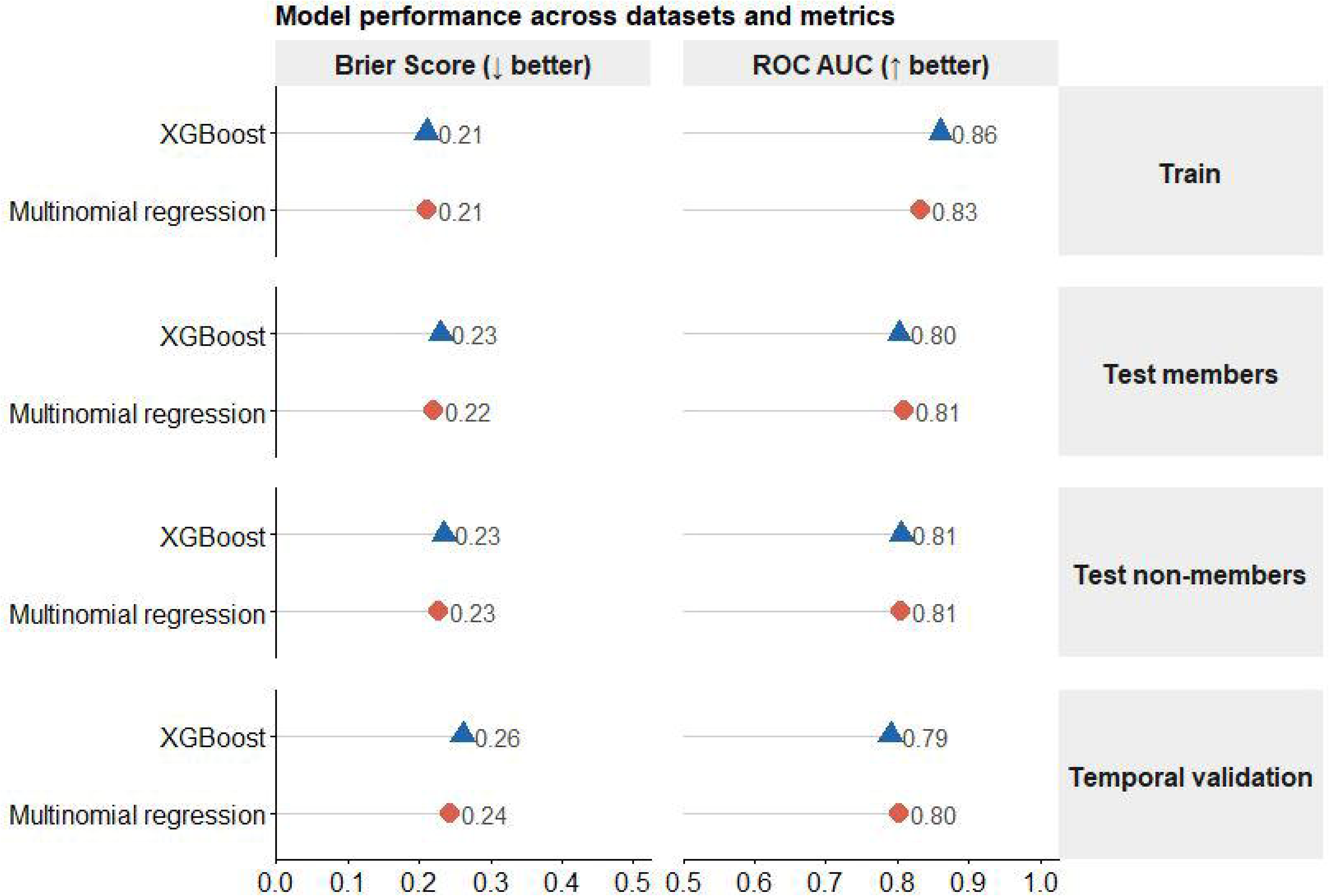
Comparative performance of models across data-sets

Performance on the non-members test set was comparable despite absent community predictors (diagnoses contributing to comorbidity indices, polypharmacy, laboratory results, allowance eligibility, and housebound status): macro-averaged AUC 0.81 (95% CI: 0.80-0.82) and Brier score 0.23.

A benchmark multinomial model using FIM alone achieved lower AUCs of 0.72 (95% CI: 0.71-0.73) on both held-out test sets, reflecting the multivariable model’s gain over the single index.

On the 2025 temporal validation set (n=1,087), the models maintained their performance, indicating stability despite potentially evolving case-mix and documentation patterns over time. The multinom reached AUC of 0.80 (95% CI: 0.78-0.82) and Brier 0.24. Small performance differences between the sets were consistent with sampling variability rather than systematic performance loss.

Table S3 depicts subgroup performance on the combined held-out sets (n=3,505). Discrimination was consistent across the orthopedic, deconditioning and neurological subgroups, but lower in ventilator-weaned patients, particularly for medical deterioration (AUC=0.61). Across socioeconomic strata, discrimination was comparable for minimal dependency and nursing-care dependency; and modestly lower for medical deterioration in the lowest stratum (AUC 0.66) but did not follow a monotonic socioeconomic gradient.

### 3.3 Alert utility

Figure 2 presents outcome-specific PPV across various alert rates on the held-out sets (n=3,505). XGBoost added no PPV over the multinomial model at any alert burden, which supports adopting the more transparent model. At a predicted absolute probability threshold of ≥60%, the multinomial model flagged 32% of patients for care-dependency and 5% for deterioration with positive predictive value of 72% and 69% respectively (Figure S3). The team deemed this precision acceptable for triggering early social-worker engagement, since a false positive (an unnecessary family conversation) costs less than a false negative (delayed social process and avoidable prolonged admission).

**Figure 2.**
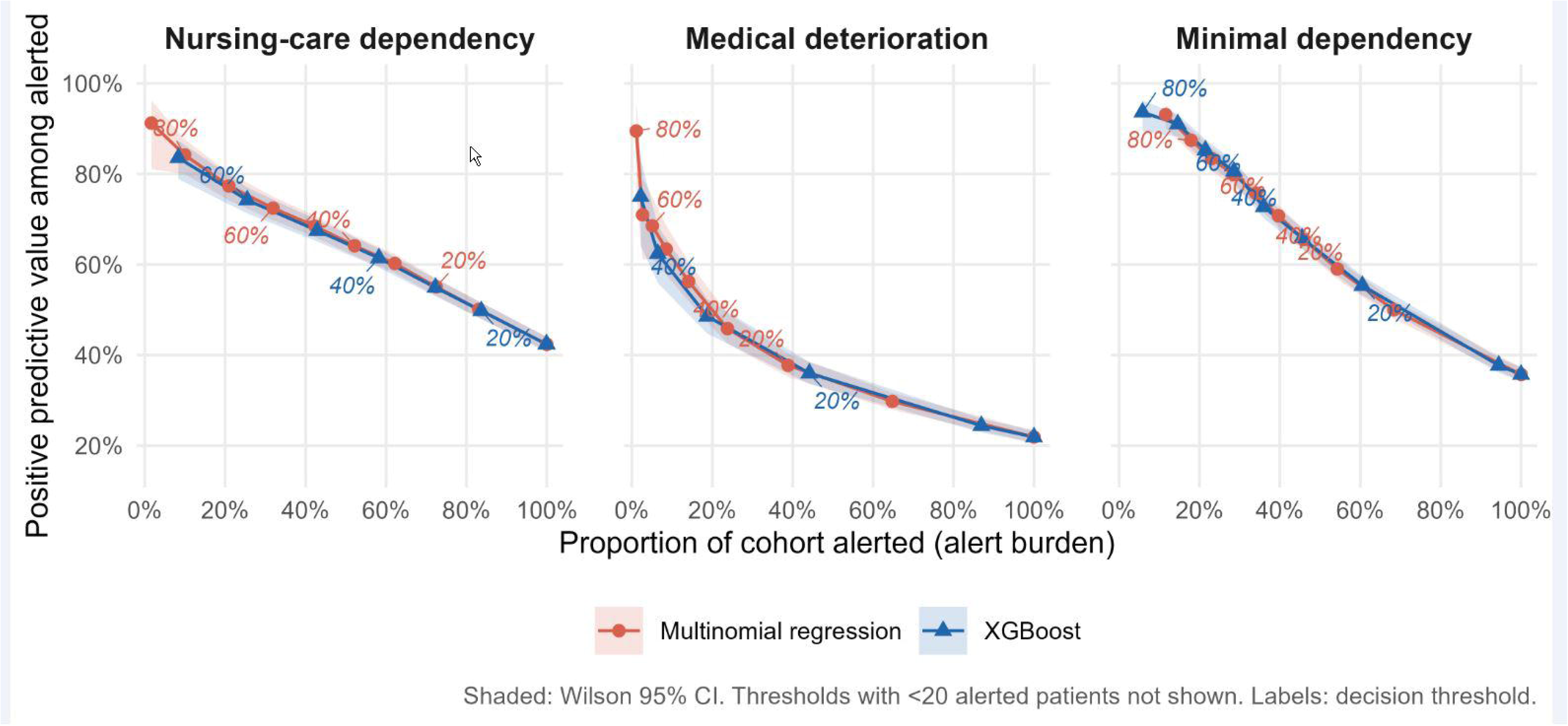
Positive predictive value (PPV) as a function of alert burden, by outcome class and model, calculated based on held-out test sets. **Point labels indicate the alert thresholds** (≥20%, ≥40%, ≥60%, ≥80%).

When dividing the held-out test sets by the most probable predicted class, the cumulative incidence of nursing-care dependency was highest among those predicted as nursing-care dependency (log-rank p<0.001; Supplementary Figure S4). The median times from index date till nursing-care dependency were 46 for nursing-care dependency (95% CI 44-48), 70 for medical deterioration (95% CI 63-78) and 82 for minimal-dependency (95% CI 78-91) days respectively.

### 3.4 Variable Importance

Table 2 depicts the overall top 15 predictors of the Multinomial model ranked by likelihood-ratio χ² statistic. First-week FIM, lab-frailty and Beiti ranked highest; FIM and Beiti were only moderately correlated (r=0.54). Per-class rankings (Figure S5) differed by outcome: laboratory frailty index ranked higher for medical deterioration than for care-dependency.

**Table 2.** Top 15 predictors ranked by likelihood ratio, Multinomial regression model.

| Characteristic | Relative importance |
|---|---|
| FIM | 100 |
| Frailty index lab | 35.0 |
| Beiti | 34.8 |
| Year | 22.9 |
| MMSE | 16.1 |
| Age | 15.5 |
| iADL | 12.5 |
| Admission reason | 9.2 |
| No. labs (first week) | 9.1 |
| Pre-admission mobility status | 7.8 |
| Population sector | 6.7 |
| Admission FIM missing | 5.3 |
| Antibiotics during admission | 5.0 |
| Premorbid motor FIM missing | 4.4 |
| Opiates during admission | 4.4 |

### 3.5 Contributors Example

The patient illustrated in Figure 3 had predicted probabilities of 76% for nursing-care dependency and 14% for medical deterioration. Nursing-care dependency odds (vs. minimal-dependency) were mainly driven by low admission FIM and premorbid motor FIM, comorbidities (high deficits counts and Elixhauser), low Beiti score and iADL; an above average MMSE was protective.

**Figure 3.**
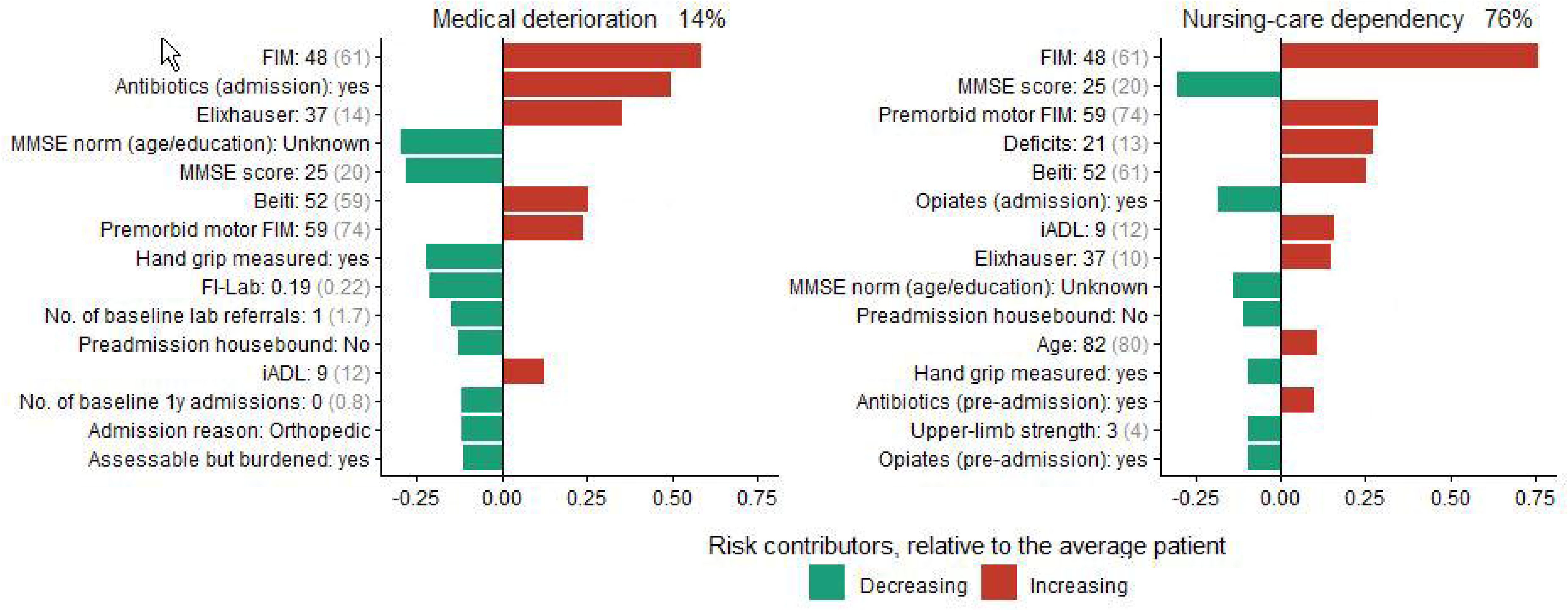
Local explanation for an illustrative patient. Each bar shows how much a predictor shifts the log-odds of that outcome relative to minimal dependency, compared with the average patient in the training cohort: red bars increase and green bars decrease these odds. Bars are sorted by absolute contribution, and the 15 largest per outcome are shown. Grey values in brackets are reference values, at which the predictor contributes nothing.

Medical deterioration odds were driven by low FIM, Beiti, antibiotics at admission and a high Elixhauser score, partly offset by a recorded hand-grip measurement and a below-average FI-Lab.

### 3.6 Misclassification drill-down

Binary discrimination was highest for separating minimal dependency from the other two outcomes (one-vs-rest AUC 0.89), and lowest for medical deterioration (one-vs-rest AUC 0.75). A multiclass AUC averages discrimination across classes and is inherently lower than a binary AUC for the most separable split. Most multiclass model misclassifications were medical deterioration cases predicted as nursing-care dependency. Missed (false-negative) deteriorations occurred later than predicted ones (mean 31 vs. 12 days), and had lower day-7 FI-Lab and better, more complete functional scores than detected deteriorations.

## 4. DISCUSSION

### 4.1 Principal Findings

Across a decade of consecutive admissions, a day-7 multiclass model integrating hospital assessments with pre-admission community data achieved good discrimination and calibration. Its actionable outcomes map onto two use cases: early social-work involvement for likely nursing-care dependency and closer monitoring for high deterioration risk.

### 4.2 Leading predictors and comparison with prior literature

The largest comparable study, a US Medicare cohort of 426,680 older adults admitted to skilled nursing facilities, likewise used a day-7 landmark and reported C-statistics of 0.79 for 6-month mortality and 0.73 for successful community discharge. ^11^ Its reliance on the federally mandated Minimum Data Set favors transportability across facilities. Our model differs in two respects. First, it yields complementary probabilities for three in-stay outcomes, rather than separate post-discharge estimates. Second, it targets the level of support required rather than discharge destination, which also reflects caregiver availability, patient preference and socioeconomic status. ^9,10^ For the closest analogous dichotomy, minimal dependency vs. all other outcomes, the AUC was 0.89, compared with 0.73 for successful destination, but differences in setting, case mix and definitions preclude direct comparison of discrimination. Another machine-learning study was restricted to patients with hip fractures in one orthopedic ward.^28^ It relied on predictors that are not routinely coded (e.g., caregiver presence, weight-bearing instructions), which limits automated prospective scoring and scale-up. It also excluded the 14% who did not complete rehabilitation, mostly owing to medical deterioration or death. Because such exclusion depends on post-admission events, that model cannot flag patients who will deteriorate. Our study covers all admission reasons and discharge outcomes and uses only automatically rather than manually extracted data.

The strongest predictor for functional independence was admission FIM; Beiti, structurally similar, ranked third. Beiti correlated strongly with FIM in its original report (r=0.81-0.86) but only modestly here (r=0.54), suggesting partially non-overlapping information.^14^ Beiti is routine in Israeli rehabilitation practice but is not, to our knowledge, internationally validated. Consistent with FI-Lab as a measure of biological dysregulation, the index demonstrated strong discrimination for medical deterioration (a primarily biological outcome) but limited discrimination for rehabilitation failure (a primarily functional outcome). This aligns with prior work suggesting FI-Lab and functional frailty indices capture complementary dimensions of vulnerability. ^15,16^

### 4.3 Value of Community Health Data Linkage

The integration of pre-admission community health data provided information not fully available in the hospital’s own records, namely chronic disease registries, medication and laboratory histories, community nurses assessments, and national insurance long-term care allowance. Although some community variables ranked highly, performance and calibration differed little between the HMO members and non-members sets. This suggests that much of the pre-admission signal is also captured by first-week hospital intake.

### 4.4 The Landmark Approach and Clinical Decision Timing

A distinctive feature is anchoring prediction to the first team meeting. By day 7, initial functional assessments, admission laboratory values, and information on early complications (delirium, pressure wounds, infections etc.) are available; an admission-day model would lack these, whereas a later landmark would reduce the lead time for social or medical interventions.

Re-scoring at each subsequent team meeting as data accrue may capture deteriorations missed at day 7, which tended to occur long after the cutoff.

### 4.5 Clinical Implementation: From Prediction to Action

Beyond the model itself, frameworks for adopting decision-support tools emphasize specifying where a model’s output enters the workflow and who acts on it. ^21^ Accordingly, we designed two use cases mapping onto existing clinical workflow.

For nursing-care dependency, early social care planning allows time to align discharge arrangements with the patient’s needs and the family’s caregiving capacity. Once rehabilitation goals are met, days spent awaiting community arrangements add no therapeutic benefit. Early planning may spare patients these waiting days and, at the institutional level, free rehabilitation beds for other awaiting admission.

By triggering social process initiation at day 7 based on the model’s prediction, the social and bureaucratic processes can run in parallel with ongoing rehabilitation rather than sequentially. Triggering it early does not preclude continued rehabilitation; it is parallel preparation, not a self-fulfilling prophecy. The mean time to social intervention was 21 days, a gap partly attributable to late initiation. An analogous analysis of the medical deterioration use-case was not possible because there is currently no documentation of the medical monitoring processes.

### 4.6 Next steps

A parallel-ward controlled trial should test whether alerts shorten time to social-work initiation and length of stay and are acceptable to clinicians. External validation in other centers is needed before wider adoption.

### 4.7 Strengths and Limitations

Study strengths include the large, unselected decade-long cohort of consecutive admissions, the multiclass outcome distinguishing between unfavorable outcomes, the landmark design anchored to the multidisciplinary meeting, the integration of hospital and community data, operationally defined clinical use cases, and a local explanation framework.

Several limitations must be acknowledged. First, this single-center study’s transportability to other settings (different populations, staffing, record quality, or outcome definitions) is unconfirmed. Second, community health data were available only for 79%, though the other HMOs’ test set showed minimal performance reduction. Third, while the outcome definition of dependency used a nationally regulated standard, its documentation in the medical record may vary across facilities. Lastly, medical deterioration had the least events relative to the number of model parameters; its local explanations should be interpreted with caution.

## 5. CONCLUSIONS

Routinely collected data identified patients at high risk of nursing-care dependency or medical deterioration. Early alerts may prompt closer monitoring, earlier discharge planning, and better preparation of families while making more efficient use of rehabilitation resources.

## Data statement

The fitted prediction model is openly available at Zenodo (https://zenodo.org/records/23649297). The underlying patient-level data is not publicly available due to patient privacy regulations and institutional data-sharing restrictions. Analysis code is available from the corresponding author on reasonable request.

## Acknowledgements

We are grateful to Dr. Noa Caspi and Shir Zecharya from Clalit Data Research Center, and Vered Assa Blitzer from Beit Rivka, whose extensive assistance with data extraction made this work possible.

## List of Abbreviations

ADL: activities of daily living
AUC: area under the receiver operating characteristic curve
BMI: body mass index
CI: confidence interval
FI-Lab: laboratory frailty index
FIM: Functional Independence Measure
HMO: health maintenance organization
iADL: instrumental activities of daily living
IQR: interquartile range
MMSE: Mini-Mental State Examination
MoH: Ministry of Health
PPV: positive predictive value
ROC: receiver operating characteristic
SHAP: SHapley Additive exPlanations
TDM: therapeutic drug monitoring
TRIPOD: Transparent Reporting of a multivariable prediction model for Individual Prognosis Or Diagnosis
TSH: thyroid-stimulating hormone
XGBoost: extreme gradient boosting

See Supplementary Material for supporting figures and tables.

## REFERENCES

1. Asiskovitch S. The Long-Term Care Insurance Program in Israel: solidarity with the elderly in a changing society. Isr J Health Policy Res. 2013;2(1):3. doi:10.1186/2045-4015-2-3

2. Foster MM, Borg DN, Houston V, et al. Planning with care complexity: Factors related to discharge delays of hospitalised people with disability. Health Soc Care Community. 2022;30(6):e4992–e5000. doi:10.1111/hsc.13912

3. Harrison JK, Walesby KE, Hamilton L, et al. Predicting discharge to institutional long-term care following acute hospitalisation: a systematic review and meta-analysis. Age Ageing. 2017;46(4):547–558. doi:10.1093/ageing/afx047

4. Kumar A, Graham JE, Resnik L, et al. Comparing Comorbidity Indices to Predict Post-Acute Rehabilitation Outcomes in Older Adults. Am J Phys Med Rehabil. 2016;95(12):889–898. doi:10.1097/PHM.0000000000000527

5. Reistetter TA, Graham JE, Deutsch A, Granger CV, Markello S, Ottenbacher KJ. Utility of functional status for classifying community versus institutional discharges after inpatient rehabilitation for stroke. Arch Phys Med Rehabil. 2010;91(3):345–350. doi:10.1016/j.apmr.2009.11.010

6. Singh I, Gallacher J, Davis K, Johansen A, Eeles E, Hubbard RE. Predictors of adverse outcomes on an acute geriatric rehabilitation ward. Age Ageing. 2012;41(2):242–246. doi:10.1093/ageing/afr179

7. Fernandes S, Bula C, Krief H, Carron PN, Seematter-Bagnoud L. Unplanned transfer to acute care during inpatient geriatric rehabilitation: incidence, risk factors, and associated short-term outcomes. BMC Geriatr. 2024;24(1):456. doi:10.1186/s12877-024-05081-3

8. Aditya BS, Sharma JC, Allen SC, Vassallo M. Predictors of a nursing home placement from a non-acute geriatric hospital. Clin Rehabil. 2003;17(1):108–113. doi:10.1191/0269215503cr567oa

9. Nguyen TA, Page A, Aggarwal A, Henke P. Social Determinants of Discharge Destination for Patients After Stroke With Low Admission FIM Instrument Scores. Arch Phys Med Rehabil. 2007;88(6):740–744. doi:10.1016/j.apmr.2007.03.011

10. Inneh IA, Clair AJ, Slover JD, Iorio R. Disparities in Discharge Destination After Lower Extremity Joint Arthroplasty: Analysis of 7924 Patients in an Urban Setting. J Arthroplasty. 2016;31(12):2700–2704. doi:10.1016/j.arth.2016.05.027

11. Deardorff WJ, Gan S, Jing B, et al. An Updated Mortality and Community Discharge Prognostic Model for Older Adults Admitted to Skilled Nursing Facilities for Post-Acute Care. J Am Med Dir Assoc. 2026;27(4). doi:10.1016/j.jamda.2025.106103

12. TRIPOD+AI statement: updated guidance for reporting clinical prediction models that use regression or machine learning methods | The BMJ. Accessed October 25, 2026. https://www.bmj.com/content/385/bmj-2023-078378

13. Choosing a Nursing Care Home. Older Adults. Accessed June 29, 2026. https://me.health.gov.il/en/older-adult/services-rights/hospitalization/nursing-hospitalization/choosing-a-facility/

14. Frenkel M, Eckstein N, Frenkel-Toledo S. Psychometric characteristics of the BEITI - an assessment scale of daily functioning. NeuroRehabilitation. 2020;47(4):387–392. doi:10.3233/NRE-203122

15. Howlett SE, Rockwood MRH, Mitnitski A, Rockwood K. Standard laboratory tests to identify older adults at increased risk of death. BMC Med. 2014;12:171. doi:10.1186/s12916-014-0171-9

16. Blodgett JM, Theou O, Howlett SE, Rockwood K. A frailty index from common clinical and laboratory tests predicts increased risk of death across the life course. GeroScience. 2017;39(4):447–455. doi:10.1007/s11357-017-9993-7

17. Quan H, Li B, Couris CM, et al. Updating and validating the Charlson comorbidity index and score for risk adjustment in hospital discharge abstracts using data from 6 countries. Am J Epidemiol. 2011;173(6):676–682. doi:10.1093/aje/kwq433

18. Moore BJ, White S, Washington R, Coenen N, Elixhauser A. Identifying Increased Risk of Readmission and In-hospital Mortality Using Hospital Administrative Data: The AHRQ Elixhauser Comorbidity Index. Med Care. 2017;55(7):698–705. doi:10.1097/MLR.0000000000000735

19. Cheng D, DuMontier C, Yildirim C, et al. Updating and Validating the U.S. Veterans Affairs Frailty Index: Transitioning From ICD-9 to ICD-10. J Gerontol A Biol Sci Med Sci. 2021;76(7):1318–1325. doi:10.1093/gerona/glab071

20. Nori H, Jenkins S, Koch P, Caruana R. InterpretML: A Unified Framework for Machine Learning Interpretability. arXiv. Preprint posted online September 19, 2019:arXiv:1909.09223. doi:10.48550/arXiv.1909.09223

21. Dagan N, Devons-Sberro S, Paz Z, et al. Evaluation of AI Solutions in Health Care Organizations — The OPTICA Tool. NEJM AI. 2024;1(9):AIcs2300269. doi:10.1056/AIcs2300269

22. Amann J, Blasimme A, Vayena E, Frey D, Madai VI, Precise4Q consortium. Explainability for artificial intelligence in healthcare: a multidisciplinary perspective. BMC Med Inform Decis Mak. 2020;20(1):310. doi:10.1186/s12911-020-01332-6

23. Tonekaboni S, Joshi S, McCradden MD, Goldenberg A. What Clinicians Want: Contextualizing Explainable Machine Learning for Clinical End Use. In: Proceedings of the 4th Machine Learning for Healthcare Conference. PMLR; 2019:359–380. Accessed October 7, 2026. https://proceedings.mlr.press/v106/tonekaboni19a.html

24. Ramspek CL, Jager KJ, Dekker FW, Zoccali C, van Diepen M. External validation of prognostic models: what, why, how, when and where? Clin Kidney J. 2020;14(1):49–58. doi:10.1093/ckj/sfaa188

25. Pate A, Riley RD, Collins GS, et al. Minimum sample size for developing a multivariable prediction model using multinomial logistic regression. Stat Methods Med Res. 2023;32(3):555–571. doi:10.1177/09622802231151220

26. Riley RD, Ensor J, Snell KIE, et al. Calculating the sample size required for developing a clinical prediction model. BMJ. 2020;368:m441. doi:10.1136/bmj.m441

27. Hand DJ, Till RJ. A Simple Generalisation of the Area Under the ROC Curve for Multiple Class Classification Problems. Mach Learn. 2001;45(2):171–186. doi:10.1023/A:1010920819831

28. Shtar G, Rokach L, Shapira B, Nissan R, Hershkovitz A. Using Machine Learning to Predict Rehabilitation Outcomes in Postacute Hip Fracture Patients. Arch Phys Med Rehabil. 2021;102(3):386–394. doi:10.1016/j.apmr.2020.08.011

