## Supplementary materials for "Early Prediction of Discharge Outcomes in Geriatric Rehabilitation: Development and Temporal Validation of a Multiclass Model"

Table S1. Additional baseline characteristics by outcome. Main cohort (2016-2024).

Table S2. Temporal test set, baseline characteristics by outcome group

Table S3. Subgroup performance

Figure S1. Study population and partition into train and evaluation sets

Figure S2. Multinomial regression outcome-specific calibration plots

Figure S3. Positive predictive value (PPV) as a function of decision threshold (alert)

Figure S4. Time from rehabilitation admission to Nursing-care dependency

Figure S5. Top 15 predictors per outcome class

**Table S1.** Additional baseline characteristics by outcome. Main cohort (2016-2024).

| **Characteristic** | **Minimal-**  **dependency**  N = 3,241 | **Medical deterioration**  N = 2,088 | **Nursing-care dependent**  N = 4,207 |
| --- | --- | --- | --- |
| Residential socioeconomic group |  |  |  |
| High | 1,359 (42) | 948 (45) | 1,853 (44) |
| Medium | 1,341 (41) | 814 (39) | 1,655 (39) |
| Low | 483 (15) | 287 (14) | 624 (15) |
| Unknown | 58 (2) | 39 (2) | 75 (2) |
| Cancer, n (%) | 1,236 (38) | 930 (45) | 1,720 (41) |
| Falls (diagnoses), n (%) | 1,642 (51) | 1,000 (48) | 2,315 (55) |
| Falls during admission, n (%) | 64 (2) | 72 (3) | 154 (4) |
| Amputation, n (%) | 302 (9) | 247 (12) | 403 (10) |
| Implants, n (%) | 78 (2) | 81 (4) | 105 (2) |
| TPN, n (%) | 35 (1) | 34 (2) | 33 (1) |
| Transfusion, n (%) | 33 (1) | 106 (5) | 75 (2) |
| Severe mental illness, n (%) | 1,037 (32) | 811 (39) | 1,873 (45) |
| Left ventricular assist device, n (%) | 1 (0) | 6 (0) | 1 (0) |
| Advanced CHF, n (%) | 205 (6) | 282 (14) | 519 (12) |
| Chronic heart failure, n (%) | 423 (13) | 546 (26) | 740 (18) |
| Tracheostomy, n (%) | 144 (4) | 123 (6) | 182 (4) |
| Chronic obstructive pulmonary disease, n (%) | 651 (20) | 477 (23) | 799 (19) |
| Palliative care, n (%) | 1 (0) | 12 (1) | 8 (0) |
| Stoma, n (%) | 62 (2) | 61 (3) | 66 (2) |
| Catheter, n (%) | 322 (10) | 576 (28) | 786 (19) |
| Zonda, n (%) | 141 (4) | 201 (10) | 270 (6) |
| Drain, n (%) | 43 (1) | 64 (3) | 34 (1) |
| Central venous catheter, n (%) | 61 (2) | 86 (4) | 70 (2) |
| Resistant Bacteria, n (%) | 112 (3) | 151 (7) | 165 (4) |
| Pressure wounds, n (%) | 114 (4) | 205 (10) | 241 (6) |
| Dialysis, n (%) | 47 (1) | 102 (5) | 66 (2) |
| Psychoactive medications preadmission, n (%) | 2,617 (81) | 1,774 (85) | 3,652 (87) |
| Opiates preadmission, n (%) | 2,310 (71) | 1,481 (71) | 2,783 (66) |
| Insulin preadmission, n (%) | 837 (26) | 702 (34) | 1,165 (28) |
| Antibiotics preadmission, n (%) | 1,248 (39) | 1,195 (57) | 1,992 (47) |
| Therapeutic drug monitoring medications preadmission, n (%) | 394 (12) | 384 (18) | 639 (15) |
| Psychoactive medications during admission, n (%) | 2,182 (67) | 1,483 (71) | 3,106 (74) |
| Therapeutic drug monitoring medications during admission, n (%) | 213 (7) | 208 (10) | 273 (6) |
| Opiates during admission, n (%) | 1,530 (47) | 795 (38) | 1,431 (34) |
| Insulin during admission, n (%) | 602 (19) | 543 (26) | 885 (21) |
| Antibiotics during admission, n (%) | 184 (6) | 377 (18) | 320 (8) |
| Prior admissions over 30 days, n (%) | 296 (9) | 395 (19) | 600 (14) |
| Hemoglobin < 8 g/L, n (%) | 25 (1) | 69 (3) | 37 (1) |
| Aphasia, n (%) | 93 (3) | 154 (7) | 367 (9) |
| Admission reason, n (%) |  |  |  |
| Orthopedic | 1,701 (52) | 834 (40) | 1,916 (46) |
| Deconditioning | 869 (27) | 661 (32) | 1,112 (26) |
| Neurologic | 494 (15) | 432 (21) | 957 (23) |
| Ventilated | 177 (5.5) | 161 (7.5) | 222 (5.3) |
| No. of baseline (1 year) inpatient admissions, n (%) | |  |  |
| 0 | 2,200 (68) | 1,101 (53) | 2,550 (61) |
| 1 | 619 (19) | 476 (23) | 896 (21) |
| 2 | 237 (7) | 235 (11) | 400 (10) |
| 3 | 87 (3) | 134 (6) | 178 (4) |
| 4 | 60 (2) | 59 (3) | 93 (2) |
| 5+ | 38 (1) | 82 (4) | 89 (2) |
| No. of laboratory tests in the first week, n (%) |  |  |  |
| 0 | 100 (3) | 53 (3) | 87 (2) |
| 1 | 1,797 (55) | 768 (37) | 2,217 (53) |
| 2 | 939 (29) | 696 (33) | 1,261 (30) |
| 3 | 290 (9) | 369 (18) | 489 (11) |
| 4 | 83 (3) | 151 (7) | 123 (3) |
| 5 | 25 (1) | 30 (1) | 36 (1) |
| 6 | 5 (0) | 17 (1) | 11 (0) |
| 7 | 2 (0) | 3 (0) | 2 (0) |
| Systolic blood pressure, n (%) |  |  |  |
| 130-139 | 581 (18) | 347 (17) | 746 (18) |
| 140 or more | 975 (30) | 569 (27) | 1,243 (30) |
| Under130 | 1,321 (41) | 947 (45) | 1,750 (42) |
| Unknown | 364 (11) | 224 (11) | 467 (11) |
| Diastolic blood pressure <60, n (%) | 467 (14) | 387 (19) | 756 (18) |
| Diastolic blood pressure>=90, n (%) | 149 (5) | 76 (4) | 155 (4) |
| Saturation, n (%) |  |  |  |
| 95+ | 2,255 (70) | 1,402 (67) | 2,908 (69) |
| Under 95 | 264 (8) | 333 (16) | 453 (11) |
| Unknown | 722 (22) | 352 (17) | 845 (20) |
| Polypharmacy, n (%) |  |  |  |
| 10+ | 1,072 (33) | 840 (40) | 1,664 (40) |
| 0-4 | 523 (16) | 243 (12) | 539 (13) |
| 5-9 | 928 (29) | 557 (27) | 1,220 (29) |
| Unknown | 720 (22) | 447 (21) | 783 (19) |
| MMSE, Median (IQR) | 26 (22 – 28) | 20 (14 – 25) | 18 (13 – 23) |
| Unknown | 145 | 302 | 258 |
| Typical MMSE, n (%) |  |  |  |
| Unknown | 644 (20) | 555 (27) | 945 (22) |
| Yes | 692 (21) | 206 (10) | 309 (7) |
| No | 1,905 (59) | 1,326 (64) | 2,952 (70) |
| iADL, n (%) | 17 (12 - 21) | 10 (5 - 17) | 8 (4 - 14) |
| Unknown | 321 | 506 | 653 |
| Beiti, Median (IQR) | 70 (59 – 83) | 53 (39 – 65) | 54 (42 – 64) |
| Unknown | 247 | 378 | 359 |
| FIM baseline, Median (IQR) | 78 (63 - 91) | 49 (39 - 62) | 51 (40 - 62) |
| Unknown | 25 | 10 | 31 |
| Frailty index lab, Median (IQR) | 0.15  (0.11- 0.19) | 0.20 (0.16-0.25) | 0.16 (0.12-0.21) |
| Unknown | 3 | 4 | 6 |

**Table S2.** Temporal test set, baseline Characteristics by Outcome Group

| Characteristic | Minimal-  dependency  n = 348 | Medical deterioration  n = 331 | Nursing-care dependent  n = 408 |
| --- | --- | --- | --- |
| Age, Median (IQR) | 76 (72 – 82) | 81 (74 – 87) | 83 (78 – 88) |
| Sex, n (%) |  |  |  |
| Female | 213 (61) | 169 (51) | 242 (59) |
| Male | 135 (39) | 162 (49) | 166 (41) |
| Strength upper limbs, n (%) |  |  |  |
| 2 | 7 (2.9) | 8 (4.9) | 14 (5.2) |
| 3 | 46 (19) | 55 (34) | 90 (33) |
| 4 | 101 (42) | 80 (49) | 124 (46) |
| 5 | 87 (36) | 19 (12) | 42 (16) |
| Unknown | 107 | 169 | 138 |
| Strength lower limbs, n (%) |  |  |  |
| 2 | 19 (7.5) | 35 (23) | 45 (17) |
| 3 | 90 (36) | 58 (38) | 106 (40) |
| 4 | 102 (40) | 51 (33) | 93 (35) |
| 5 | 42 (17) | 9 (5.9) | 20 (7.6) |
| Unknown | 95 | 178 | 144 |
| Hand grip, Median (IQR) | 16 (12 – 22) | 12 (8 – 19) | 12 (8 – 17) |
| Unknown | 167 | 196 | 204 |
| Nursing-care allowance (hospital data), n (%) | 94 (27) | 137 (41) | 225 (55) |
| Nursing-care eligibility level (community data), n (%) | |  |  |
| 0 | 256 (74) | 203 (61) | 188 (46) |
| 1 | 15 (4.3) | 12 (3.6) | 9 (2.2) |
| 2 | 25 (7.2) | 15 (4.5) | 25 (6.1) |
| 3 | 29 (8.3) | 32 (9.7) | 46 (11) |
| 4 | 13 (3.7) | 29 (8.8) | 40 (9.8) |
| 5 | 9 (2.6) | 10 (3.0) | 40 (9.8) |
| 6 | 1 (0.3) | 30 (9.1) | 60 (15) |
| Alcohol use disorder, n (%) | 10 (2.9) | 7 (2.1) | 5 (1.2) |
| Housebound, n (%) | 6 (1.7) | 32 (9.7) | 52 (13) |
| Pre-admission Mobility status, n (%) |  |  |  |
| Mobile | 338 (97) | 276 (83) | 311 (76) |
| Housebound | 0 (0) | 9 (2.7) | 10 (2.5) |
| Bedridden | 1 (0.3) | 2 (0.6) | 1 (0.2) |
| Mobile with assistance | 9 (2.6) | 44 (13) | 86 (21) |
| Occupational profile, Median (IQR) | 79 (76 – 89) | 75 (65 – 78) | 71 (64 – 78) |
| Unknown | 83 | 107 | 92 |
| Smoking, n (%) |  |  |  |
| never | 135 (39) | 131 (40) | 196 (48) |
| current | 29 (8.3) | 22 (6.6) | 21 (5.1) |
| past | 52 (15) | 34 (10) | 59 (14) |
| unknown | 132 (38) | 144 (44) | 132 (32) |
| Charlson comorbidity index, Median (IQR) | 3 (1 – 5) | 4 (2 – 6) | 4 (2 – 7) |
| VA-Frailty index deficits, Median (IQR) | 9.0 (6.0 – 13.0) | 11.0 (7.0 – 15.0) | 13.0 (8.0 – 16.0) |
| Elixhauser score, Median (IQR) | 11 (3 – 17) | 15 (7 – 24) | 15 (6 – 23) |
| Delirium screening, n (%) |  |  |  |
| No delirium | 229 (66) | 186 (56) | 271 (66) |
| Delirium | 0 (0) | 7 (2.1) | 5 (1.2) |
| Unscreened | 119 (34) | 138 (42) | 132 (32) |
| Delirium (diagnoses), n (%) | 13 (3.7) | 27 (8.2) | 32 (7.8) |
| Population sector, n (%) |  |  |  |
| General Jewish population | 298 (86) | 276 (83) | 355 (87) |
| Ultra-orthodox Jewish | 21 (6.0) | 21 (6.3) | 19 (4.7) |
| Immigrants from the former Soviet Union | 23 (6.6) | 24 (7.3) | 26 (6.4) |
| Arab | 6 (1.7) | 10 (3.0) | 8 (2.0) |
| year, n (%) |  |  |  |
| 2024 | 25 (7.2) | 30 (9.1) | 39 (9.6) |
| 2025 | 323 (93) | 301 (91) | 369 (90) |
| BMI, n (%) | 26.6 (23.3 – 30.4) | 26.5 (23.5 – 29.6) | 26.0 (22.9 – 29.1) |
| unknown | 2 | 1 | 2 |
| Lonely, n (%) | 34 (9.8) | 30 (9.1) | 65 (16) |
| Residential socioeconomic group |  |  |  |
| High | 161 (46) | 134 (40) | 201 (49) |
| Medium | 128 (37) | 133 (40) | 155 (38) |
| Low | 46 (13) | 57 (17) | 43 (11) |
| Unknown | 13 (4) | 7 (2) | 9 (2) |
| Cancer, n (%) | 141 (41) | 150 (45) | 182 (45) |
| Falls (diagnoses), n (%) | 163 (47) | 157 (47) | 251 (62) |
| Falls during admission, n (%) | 11 (3.2) | 14 (4.2) | 15 (3.7) |
| Amputation, n (%) | 29 (8.3) | 38 (11) | 29 (7.1) |
| Implants, n (%) | 5 (1.4) | 16 (4.8) | 13 (3.2) |
| TPN, n (%) | 5 (1.4) | 7 (2.1) | 4 (1.0) |
| Transfusion, n (%) | 1 (0.3) | 16 (4.8) | 0 (0) |
| Severe mental illness, n (%) | 100 (29) | 97 (29) | 153 (38) |
| Left ventricular assist device, n (%) | 0 (0) | 0 (0) | 0 (0) |
| Advanced CHF, n (%) | 25 (7.2) | 36 (11) | 48 (12) |
| Chronic heart failure, n (%) | 46 (13) | 89 (27) | 102 (25) |
| Tracheostomy, n (%) | 21 (6.0) | 20 (6.0) | 13 (3.2) |
| Chronic obstructive pulmonary disease, n (%) | 58 (17) | 61 (18) | 74 (18) |
| Palliative care, n (%) | 2 (0.6) | 2 (0.6) | 5 (1.2) |
| Stoma, n (%) | 12 (3.4) | 9 (2.7) | 8 (2.0) |
| Catheter, n (%) | 34 (9.8) | 93 (28) | 68 (17) |
| Zonda, n (%) | 20 (5.7) | 40 (12) | 11 (2.7) |
| Drain, n (%) | 6 (1.7) | 9 (2.7) | 2 (0.5) |
| Central venous catheter, n (%) | 10 (2.9) | 12 (3.6) | 5 (1.2) |
| Resistant Bacteria, n (%) | 27 (7.8) | 40 (12) | 21 (5.1) |
| Pressure wounds, n (%) | 12 (3.4) | 25 (7.6) | 18 (4.4) |
| Dialysis, n (%) | 4 (1.1) | 9 (2.7) | 6 (1.5) |
| Psychoactive medications preadmission, n (%) | 208 (60) | 232 (70) | 307 (75) |
| Opiates preadmission, n (%) | 265 (76) | 218 (66) | 307 (75) |
| Insulin preadmission, n (%) | 89 (26) | 113 (34) | 112 (27) |
| Antibiotics preadmission, n (%) | 195 (56) | 195 (59) | 226 (55) |
| Therapeutic drug monitoring medications preadmission, n (%) | 20 (5.7) | 38 (11) | 28 (6.9) |
| Psychoactive medications during admission, n (%) | 148 (43) | 192 (58) | 245 (60) |
| Therapeutic drug monitoring medications during admission, n (%) | 15 (4.3) | 17 (5.1) | 8 (2.0) |
| Opiates during admission, n (%) | 171 (49) | 121 (37) | 176 (43) |
| Insulin during admission, n (%) | 75 (22) | 106 (32) | 90 (22) |
| Antibiotics during admission, n (%) | 34 (9.8) | 69 (21) | 38 (9.3) |
| Prior admissions over 30 days, n (%) | 25 (7.2) | 28 (8.5) | 36 (8.8) |
| Hemoglobin < 8 g/L, n (%) | 1 (0.3) | 2 (0.6) | 1 (0.2) |
| Aphasia, n (%) | 13 (3.7) | 27 (8.2) | 24 (5.9) |
| Admission reason, n (%) |  |  |  |
| Orthopedic | 173 (50) | 124 (37) | 224 (55) |
| Deconditioning | 102 (29) | 94 (28) | 101 (25) |
| Neurologic | 54 (16) | 89 (27) | 70 (17) |
| Ventilated | 19 (5.5) | 24 (7.3) | 13 (3.2) |
| No. baseline (1 year) inpatient admissions,  Median (IQR) | 0 (0 – 1) | 0 (0 – 1) | 0 (0 – 1) |
| No. of laboratory tests (first week), n (%) |  |  |  |
| 0 | 13 (3.7) | 5 (1.5) | 5 (1.2) |
| 1 | 136 (39) | 103 (31) | 151 (37) |
| 2 | 129 (37) | 105 (32) | 144 (35) |
| 3 | 53 (15) | 57 (17) | 75 (18) |
| 4 | 11 (3.2) | 45 (14) | 25 (6.1) |
| 5 | 5 (1.4) | 15 (4.5) | 8 (2.0) |
| 6 | 0 (0) | 1 (0.3) | 0 (0) |
| 7 | 1 (0.3) | 0 (0) | 0 (0) |
| Systolic blood pressure, n (%) |  |  |  |
| 130-139 | 73 (21) | 47 (14) | 81 (20) |
| 140 or more | 75 (22) | 84 (25) | 100 (25) |
| Under130 | 195 (56) | 198 (60) | 224 (55) |
| Unknown | 5 (1.4) | 2 (0.6) | 3 (0.7) |
| Diastolic blood pressure, n (%) |  |  |  |
| 60-89 | 296 (85) | 273 (82) | 359 (88) |
| Under60 | 40 (11) | 49 (15) | 43 (11) |
| 90 or more | 7 (2.0) | 7 (2.1) | 3 (0.7) |
| Unknown | 5 (1.4) | 2 (0.6) | 3 (0.7) |
| Saturation, n (%) |  |  |  |
| 95+ | 284 (82) | 231 (70) | 313 (77) |
| Under 95 | 28 (8.0) | 77 (23) | 55 (13) |
| Unknown | 36 (10) | 23 (6.9) | 40 (9.8) |
| Polypharmacy, n (%) |  |  |  |
| 10+ | 128 (37) | 144 (44) | 213 (52) |
| 0-4 | 53 (15) | 25 (7.6) | 28 (6.9) |
| 5-9 | 78 (22) | 48 (15) | 73 (18) |
| Unknown | 89 (26) | 114 (34) | 94 (23) |
| MMSE, Median (IQR) | 26.0 (23.0 – 28.0) | 21.0 (15.0 – 24.5) | 21.0 (16.0 – 24.3) |
| Unknown | 29 | 96 | 48 |
| Typical MMSE, n (%) |  |  |  |
| Unknown | 36 (10) | 101 (31) | 57 (14) |
| Yes | 79 (23) | 26 (7.9) | 37 (9.1) |
| No | 233 (67) | 204 (62) | 314 (77) |
| iADL, n (%) | 18 (14 – 22) | 10 (5 – 18) | 9 (4 – 15) |
| Unknown | 57 | 110 | 97 |
| Beiti, Median (IQR) | 71 (59 – 83) | 53 (38 – 66) | 57 (48 – 66) |
| Unknown | 20 | 72 | 28 |
| FIM baseline, Median (IQR) | 72 (56 – 84) | 46 (37 – 58) | 52 (43 – 62) |
| Unknown | 0 | 1 | 0 |
| Frailty index lab, Median (IQR) | 0.15 (0.11 – 0.19) | 0.20 (0.15 – 0.24) | 0.17 (0.13 – 0.20) |

**Table S3- Model performance across population subgroups in the held-out test sets**

Outcome-specific area under the receiver operating characteristic curve (AUC) is evaluated one-versus-rest. Due to inherent non-collapsibility, the pooled AUC is not an average of the subgroup values. Observed/Expected: ratio of observed to expected (sum of predicted probabilities) events, with exact Poisson 95% CI.

|  |  | **Nursing-care dependency** | | |  | **Medical deterioration** | | |
| --- | --- | --- | --- | --- | --- | --- | --- | --- |
| **Subgroup** | | **Events, n** | **AUC (95% CI)** | **Observed/Expected (95% CI)** |  | **Events, n** | **AUC (95% CI)** | **Observed/Expected (95% CI)** |
| **Overall** | (n=3,505) | 1,485 | 0.81 (0.80–0.82) | 1.00 (0.95–1.05) |  | 768 | 0.75 (0.73–0.77) | 1.06 (0.99–1.14) |
| **Sector** | General (n=3,205) | 1,362 | 0.81 (0.80–0.83) | 1.00 (0.95–1.06) |  | 707 | 0.75 (0.73–0.77) | 1.06 (0.99–1.14) |
|  | Ultra-Orthodox (n=134) | 57 | 0.79 (0.71–0.86) | 0.86 (0.65–1.11) |  | 28 | 0.68 (0.58–0.78) | 1.32 (0.88–1.91) |
|  | Immigrants (n=166) | 66 | 0.78 (0.71–0.85) | 1.04 (0.80–1.32) |  | 33 | 0.73 (0.64–0.82) | 0.94 (0.65–1.32) |
| **Admission reason** | Deconditioning (n=962) | 391 | 0.80 (0.77–0.83) | 0.99 (0.89–1.09) |  | 236 | 0.73 (0.70–0.77) | 1.07 (0.94–1.22) |
|  | Neurological (n=697) | 349 | 0.79 (0.75–0.82) | 1.03 (0.92–1.14) |  | 165 | 0.73 (0.68–0.77) | 1.11 (0.95–1.29) |
|  | Orthopedic (n=1,621) | 652 | 0.83 (0.81–0.85) | 0.97 (0.90–1.05) |  | 312 | 0.78 (0.75–0.81) | 1.07 (0.96–1.20) |
|  | Post-ventilated (n=225) | 93 | 0.73 (0.67–0.80) | 1.08 (0.87–1.32) |  | 55 | 0.61 (0.53–0.70) | 0.89 (0.67–1.16) |
| **Socioeconomic status** | High / very high (n=1,517) | 640 | 0.80 (0.78–0.82) | 0.99 (0.91–1.07) |  | 353 | 0.74 (0.72–0.77) | 1.11 (1.00–1.23) |
|  | Medium (n=1,428) | 605 | 0.82 (0.79–0.84) | 1.01 (0.93–1.09) |  | 300 | 0.78 (0.75–0.81) | 1.02 (0.90–1.14) |
|  | Low / very low (n=488) | 207 | 0.82 (0.78–0.86) | 0.97 (0.84–1.11) |  | 102 | 0.66 (0.60–0.72) | 1.12 (0.91–1.36) |
|  | Not recorded (n=72) | 33 | 0.80 (0.70–0.90) | 1.22 (0.84–1.71) |  | 13 | 0.76 (0.59–0.92) | 0.79 (0.42–1.36) |

**Figure S1.** Study population and partition into train and evaluation sets

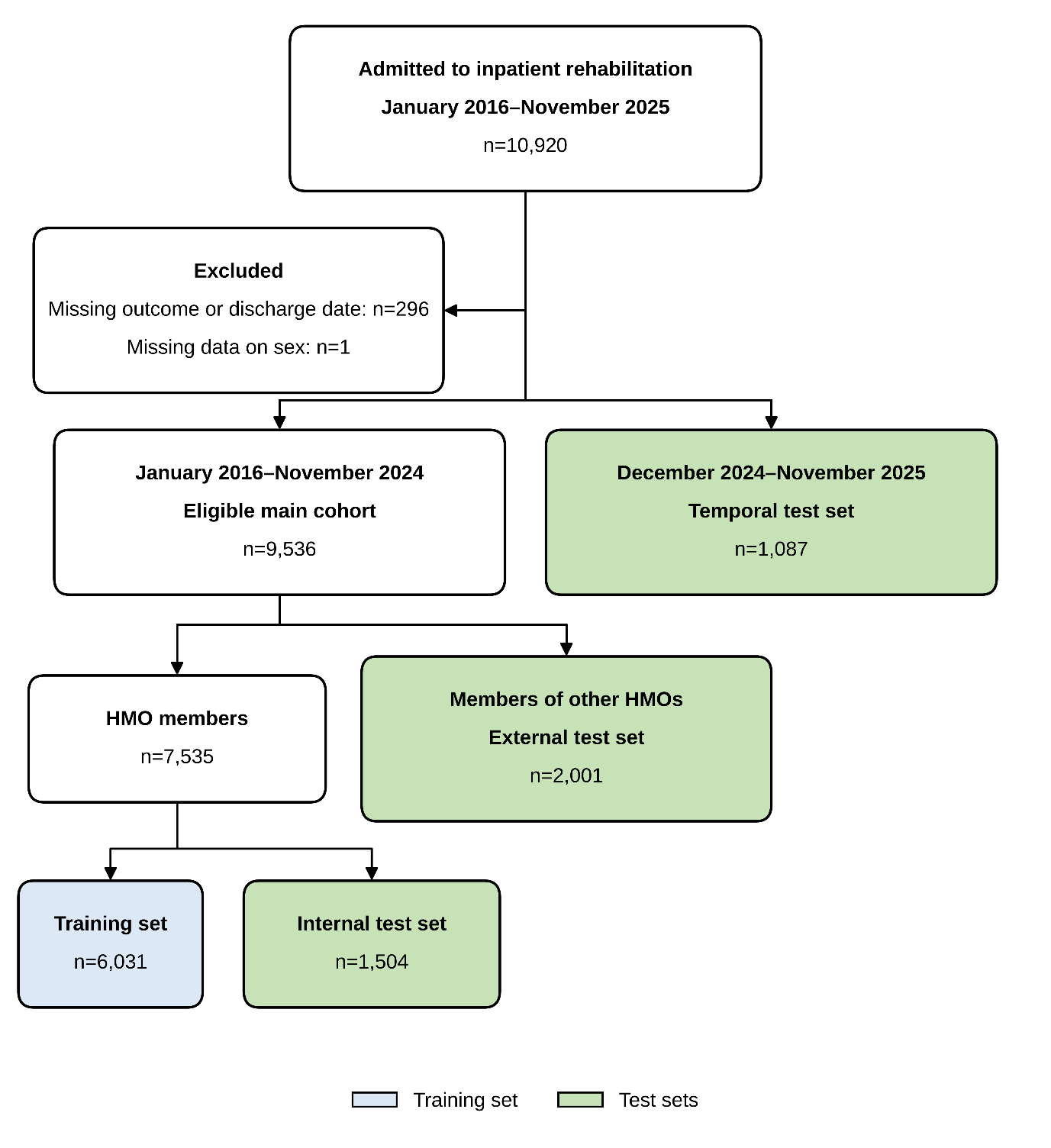

**Figure S2.** Model specific and outcome-specific calibration plots based on held-out test sets. For each outcome, points show the observed outcome rate against the mean predicted probability within deciles of predicted probability.

Dashed: perfect calibration (intercept zero, slope 1). Solid: empirical calibration (LOESS smoothed). Shades: 95% CI.

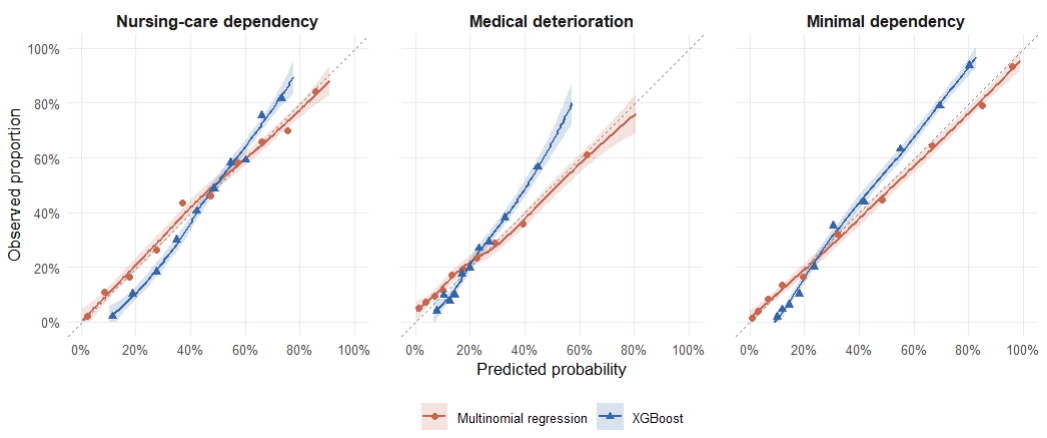

**Figure S3**. Model specific and outcome specific positive predictive value (PPV) as a function of decision threshold (alert) based on held-out test sets. At each threshold t, PPV (y axis) is computed cumulatively among all patients with predicted class probability ≥ t (x-axis). **Point labels indicate the rate of alerted patients** at selected thresholds (20%, 40%, 60%, 80%). Shades: 95% CI. Thresholds <20 patients are not shown.

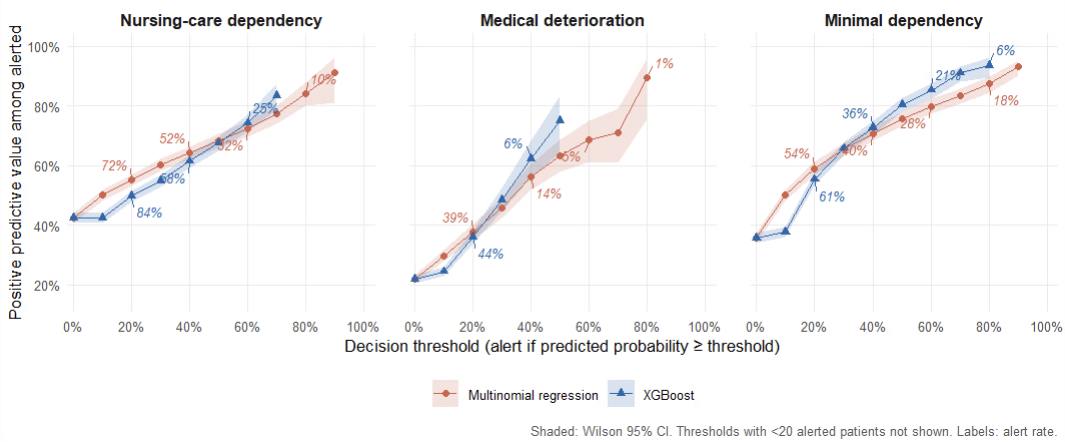

**Figure S4**. Time from rehabilitation admission to Nursing-care dependency based on the held-out test sets. Patients are stratified by the most probable predicted class.
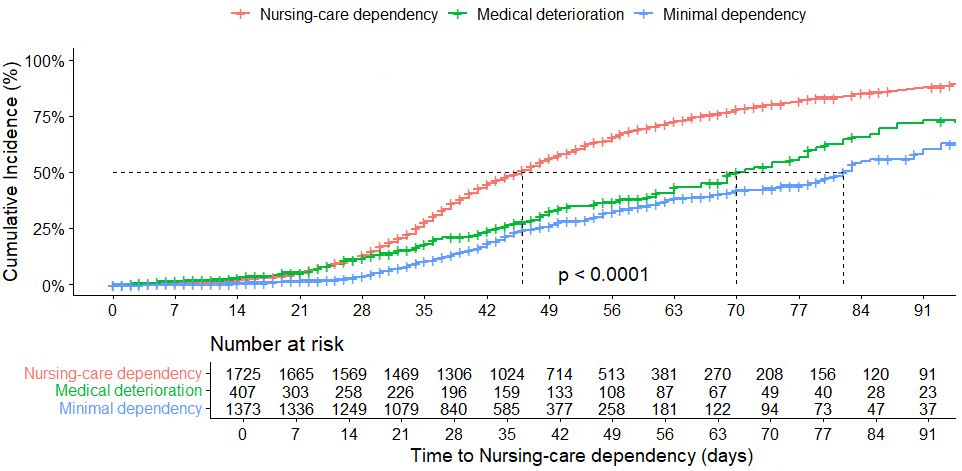

**Figure S5**. Top 15 predictors per outcome class, multinomial model. Ranked by joint Wald chi-squared statistic of each predictor's term minus its degrees of freedom (to avoid favoring multi-term predictors). Larger values indicate stronger statistical evidence for an association.

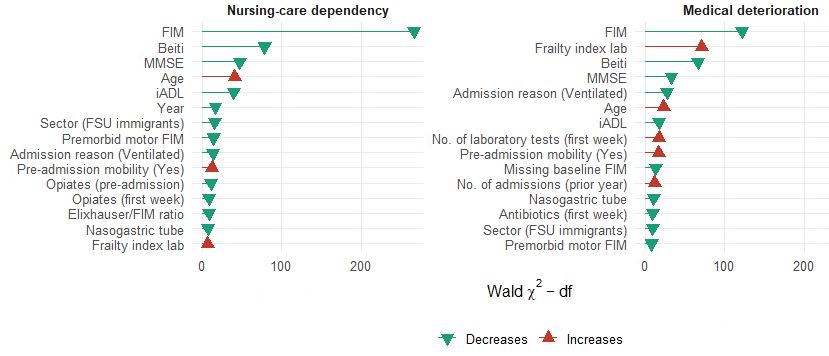
